# The Combination of Neuropsychiatric Symptoms and Blood-Based Biomarkers Enhances Early Detection of Mild Cognitive Impairment

**DOI:** 10.1101/2024.12.08.24318690

**Authors:** Yi Jin Leow, Zahinoor Ismail, Seyed Ehsan Saffari, Gurveen Kaur Sandhu, Pricilia Tanoto, Faith Phemie Hui En Lee, Smriti Ghildiyal, Shan Yao Liew, Gursimar Bhalla, Sim Xin Ying, Adnan Azam Mohammed, Ashwati Vipin, Chao Dang, Nagaendran Kandiah

## Abstract

**Background:** Combing behavioral assessments with blood-based biomarkers (BBM) may improve detection of Mild Cognitive Impairment (MCI) linked to early-stage neurodegenerative disease. Neuropsychiatric symptoms (NPS) often precede or accompany cognitive decline and provide observable behavioral signals, while BBM reflect underlying neuropathological changes. We investigated if integrating biological (plasma biomarkers) and behavioral (NPS) measures improves differentiation of MCI from cognitively normal (CN) individuals in a multi-ethnic Southeast Asian cohort— an underrepresented population in dementia research.

**Methods:** This cross-sectional analysis included 678 community-dwelling adults (mean age 59.2±11.0, 60.5% female) from the Biomarkers and Cognition Study, Singapore (BIOCIS), comprising participants recruited from the community, at Dementia Research Centre (Singapore) from February 2022 to March 2024. Participants underwent behavioral assessments using the Mild Behavioral Impairment Checklist (MBI-C) and the Depression, Anxiety, and Stress Scales (DASS). Plasma biomarkers measured were amyloid-beta (Aβ40, Aβ42), phosphorylated tau (p-tau181), neurofilament light (NfL), and glial fibrillary acidic protein (GFAP). Logistic regression and receiver operating characteristic (ROC) analyses evaluated the discriminative power of NPS, BBM, and their combination for identifying MCI risk.

**Results:** MBI-C total scores and subdomains (Mood, Interest, Control) and plasma biomarkers (Aβ40, NfL, GFAP) were significantly elevated in MCI compared to CN participants. Multivariate analysis showed elevated plasma GFAP (OR=3.64, 95% CI:1.96–6.75, p<0.001) and higher MBI-C Mood scores (OR=2.61, 95% CI:1.54–4.44, p<0.001) as the variables most associated with MCI. The combined model integrating NPS and BBM achieved a higher discriminative ability (AUC = 0.786) for MCI than models using NPS (AUC = 0.593) or BBM (AUC = 0.697) alone. The integrated model yielded 64.7% sensitivity and 84.9% specificity for distinguishing MCI from CN, outperforming single-domain approaches.

**Conclusions:** Integrating biological and behavioral markers improves identification of individuals with early cognitive impairment. Notably, GFAP-driven neuroinflammation and mood disturbances emerged as key features of prodromal dementia, highlighting astrocytic activation and affective changes as promising biomarkers and early intervention targets. This dual-domain, multimodal framework offers translational potential for earlier detection, risk stratification, and timely intervention for Alzheimer’s disease and other dementias.

## Introduction

As dementia therapeutics advance, the responsibility to leverage biomarkers for earlier and more accurate identification of cognitive impairment secondary to neurodegenerative disease (NDD) has grown accordingly. Advances in biomarker research are rapidly broadening the arsenal of minimally invasive tools available to detect cognitive impairment at the earliest and most preclinical stages^1,2^. In particular, plasma-derived and behavioral-biomarkers are increasing shown to play a crucial role for the early detection of NDD-related cognitive impairment, offer accessible and complementary perspectives—capturing both underlying pathological processes and clinically observable symptoms of cognitive decline. Such biomarkers can augment traditional cognitive assessments, positron emission tomography (PET), and structural neuroimaging.

Blood-based biomarkers(BBM) are transforming Alzheimer’s disease(AD) diagnosis by providing a minimally invasive, scalable, and accessible alternative to traditional cerebrospinal fluid(CSF) and positron emission tomography(PET) measures^3–7^. Recent research shows that BBM achieve high accuracy in predicting clinical decline in individuals with mild cognitive impairment(MCI), highlighting their potential as early indicators^8,9^. A Swedish study confirmed that BBM assays targeting amyloid-beta and phosphorylated tau achieve diagnostic accuracy comparable to CSF and PET, with predictive values exceeding 90%, demonstrating their clinical utility in various care settings^10^. Regulatory advancements have also gained momentum, exemplified by the emergence of CLIA-cleared assays, including C2N’s PrecivityAD and Quest’s AD-Detect ^11,12^, further supporting the integration of BBM into clinical practice. Compared with lumbar puncture or PET, BBM offer substantial practical advantages, notably improved scalability, reduced invasiveness, and enhanced patient compliance, particularly beneficial in resource-limited settings^13^.

Building on this clinical promise, the Amyloid–Tau–Neurodegeneration (ATN) framework offers a mechanistic scaffold for interpreting BBM signals. Within ATN, plasma amyloid β 42 (Aβ42), amyloid β 40 (Aβ40), and the Aβ42/Aβ40 ratio quantify cerebral plaque burden ^14,15^. For tau pathology, phosphorylated Tau 181(p-tau181) serves as a marker for neurofibrillary tangles associated with AD progression ^16–18^. Lu et al. ^19^ demonstrated that plasma p-tau181 strongly correlates with tau-PET imaging, accurately distinguishing Aβ-positive from Aβ-negative individuals, underscoring its scalability as a biomarker of tau pathology. Additionally, Neurofilament light(NfL) provides insights into neuroaxonal damage, while Glial fibrillary acidic protein (GFAP) is associated with reactive astrogliosis and neuroinflammation^20–22^. Multiplex ATN biomarker panels have increasingly been validated as sensitive predictors of cognitive decline and neurodegeneration^23^.

Genetic factors, particularly the apolipoprotein E ε4 (APOE4) allele, further modulate biomarker trajectories. APOE4 remains the strongest common risk variant for late-onset AD, strongly associated with accelerated amyloid deposition^24,25^. Its influence, however, is likely ancestry dependent: Southeast Asian populations exhibit a lower prevalence of APOE4 compared to Western populations ^26^, potentially explaining differences in amyloid positivity, cognitive decline rates, and dementia types observed between these groups ^27^. Recent population genomics work also shows that APOE4 effects vary by age and sex across ancestries, nuances that are increasingly considered in trial design ^28^.

Recently, the European Medicines Agency, stipulated that lecanemab could not be prescribed in APOE4 homozygotes, further supporting its importance ^29^. Collectively, BBM and genetic profiles provide comprehensive insights into neurodegeneration, enhancing understanding of AD progression.

Complementing biochemical markers, behavioral biomarkers capture early clinical changes often preceding cognitive impairment in AD and related dementias^30,31^. Neuropsychiatric symptoms(NPS) are non-cognitive, behavioral, or psychiatric symptoms that often accompany or precede cognitive decline^32^, including the subset know as Mild Behavioral Impairment(MBI). MBI, defined by the later-life onset of persistent behavioral change, has emerged as a reliable predictor of cognitive decline and incident dementia^33–35^. The Mild Behavioral Impairment Checklist(MBI-C) was developed explicitly to measure MBI, and comprises five domains including decreased motivation, emotional dysregulation, impulse dyscontrol, social inappropriateness, and abnormal perception or thoughts^36^.

The MBI-C effectively distinguishes NPS driven by NDD, from NPS with psychiatric or stress-related etiologies, thereby enhancing dementia risk assessment^37–39^. Complementarily, the Depression, Anxiety, and Stress Scales(DASS) ^40^ provide a comprehensive evaluation of mood symptoms in dementia increasingly recognized as prodromal indicators of cognitive decline^41,42^. Together, the MBI-C and DASS capture a broad spectrum of behavioural and psychological changes that correlates with cerebrospinal fluid(CSF)^43^, plasma biomarkers^44,45^, and cognitive impairment^46,47^.

Emerging evidence suggests behavioral changes have biological underpinnings^48^, reinforcing NPS as potential clinical harbingers of neuropathology. However, evidence linking NPS with specific BBM remains mixed. For instance, in a large population-based study, no difference in AD risk was observed between APOE4 carriers and non-carriers among Americans without baseline depression^49^. For amyloid-related biomarkers, amyloid-beta has been more extensively studied in relation to behavioral symptoms, particularly depression^50^. A study in Vienna reported that higher baseline plasma Aβ42 levels predicted incident depression and conversion to AD over five years^51^. Similarly, the Rotterdam Study reported cross-sectional links between high Aβ40 and depressive symptoms in prodromal dementia, but longitudinally associated lower Aβ40 and Aβ42 with increased depression risk in elderly individuals without dementia, indicating a complex role of Aβ peptides in the etiology of depression^52^. For tau biomarkers, elevated plasma p-tau181 has been associated with NPS such as appetite changes and disinhibition^53^. Higher baseline plasma p-tau181 was also shown to predict the NPS over 5 years^54^. Regarding neurodegeneration biomarkers, NfL has shown promise as a marker for multiple NPS, including aberrant motor behavior, anxiety, sleep disturbances and euphoria^55^. GFAP, a marker of astrocytic activation and neurodegeneration, has shown mixed associations in prior studies—linked to depression in some^56^ but not significantly associated with NPS in others^57^.

Specifically focusing on MBI, recent studies link its domains more consistently to ATN biomarkers. For amyloid-related biomarkers, overall MBI status and the subdomain of affective dysregulation demonstrated associations with amyloid-beta^58^. In terms of tau pathology, recent findings by Ghahremani^44^ indicate MBI correlates with elevated plasma p-tau181 levels and a nearly fourfold increased risk of dementia, with higher baseline and longitudinal increases in p-tau181 in individuals with MBI compared to those without MBI. Additionally, Gonzalez Bautista^45^ found that the MBI domain of abnormal perception was associated with steeper increases in plasma p-tau181, implicating tau pathology in the behavioral changes observed during preclinical and prodromal AD. In neurodegeneration, findings from the Alzheimer’s Disease Neuroimaging Initiative(ADNI) cohort revealed that in preclinical and prodromal samples, MBI and its interaction with time predict NfL changes, indicating MBI as an early marker of accelerated neurodegeneration and cognitive decline^59^. Collectively, these studies illustrate MBI’s value as an early marker of AD pathology and dementia risk.

Given that BBM and NPS each uniquely reflect the disease process—one capturing neuropathology, the other clinical behavior—their integration could significantly enhance early cognitive impairment detection. BBM alone may lack clinical specificity, while isolated NPS can result from psychiatric disorders or aging. Combining both domains may identify individuals where biological pathology aligns closely with clinical symptomatology, characterizing prodromal dementia more precisely than single-domain approaches. Indeed, combined biomarker and symptom positivity has previously demonstrated superior predictive accuracy for dementia progression.

Despite theoretical appeal, comprehensive studies explicitly evaluating the combined predictive utility of NPS and BBM for MCI detection remain scarce, as prior research assessed these domains separately. In the underrepresented, multi-ethnic Southeast Asian population, genetic, cultural, and lifestyle factors may uniquely shape both biomarker profiles and the expression of NPS, highlighting the importance of integrated approaches. Thus, this study examines whether combining NPS, as measured by the MBI-C and DASS, with blood biomarkers (GFAP, NfL, p-tau181, Aβ42/40), improves the differentiation of individuals with MCI from cognitively normal(CN) individuals in a Southeast Asian population. We further explore relationships between NPS and BBM, aiming to clarify mechanistic links between behavior and neurobiology during the prodromal dementia stage.

## Methods

### Participants

This cross-sectional study recruited community-dwelling participants in Singapore from the Biomarkers and Cognition Study (BIOCIS). Data was collected from February 2022 to March 2024. Individuals were eligible if they were aged 30 to 95 years, had intact mental capacity, and were literate in English or Mandarin. Exclusion criteria encompassed serious neurological, psychiatric, or systemic illnesses, psychotic disorders, major depressive disorder, alcoholism, or drug dependency within the past two years. Additional details on study design and methodology can be found in the previously published BIOCIS protocol paper ^60^.

### Neuropsychological Test Battery

Participants underwent a comprehensive neuropsychological assessment administered by trained research personnel in either English or Mandarin. Global cognition was measured using the Montreal Cognitive Assessment (MoCA) and the Visual Cognitive Assessment Test (VCAT). Learning and memory were evaluated with the Rey Auditory Verbal Learning Test (RAVLT) and the Free and Cued Selective Reminding Test (FCSRT), while working memory was assessed using the Digit Span Backward subtest from the WAIS-IV. Executive function was tested with the Test of Practical Judgment (TOPJ), Color Trails Test 2 (CT-2), and WAIS-IV Block Design. Processing speed was examined using Color Trails Test 1 (CT-1), CT-2, and WAIS-IV Coding. Language was assessed with a semantic fluency task (animals), visuospatial abilities with the WAIS-IV Block Design and Rey Complex Figure Test (RCFT), and attention with the WAIS-IV Digit Span Forward subtest.

### Diagnostic Classification

Following the completion of the neuropsychological assessments, participants were categorized as either Cognitively Normal (CN) or having Mild Cognitive Impairment (MCI) based on Peterson’s^61^ criteria and National Institute on Aging-Alzheimer’s Association(NIA-AA) guidelines^62^. CN participants demonstrated standard performance across cognitive assessments without subjective memory complaints as assessed by the Subjective Memory Complaints Questionnaire (SMCQ).

Participants classified as MCI had cognitive test scores exceeding 1.5 standard deviations below the mean of the CN group and reported subjective memory complaints.

### Behavioral profiles and Neuropsychiatric symptoms

Self-reported questionnaires, including the Mild Behavioral Impairment Checklist (MBI-C) and the Depression, Anxiety, and Stress Scale (DASS), were administered to participants. The MBI-C^36^ evaluates MBI across five NPS domains: decreased motivation (Interest), emotional dysregulation (Mood), impulse dyscontrol (Control), social inappropriateness (Social), and abnormal perception or thought content (Beliefs). Symptom endorsement requires a persistent change from the individual’s longstanding behavior lasting at least six months. The DASS^40^ measures the severity of depressive, anxious, and stress-related symptoms experienced over the preceding week. For both the MBI-C and DASS, higher scores reflect greater symptom severity.

### Blood based biomarkers

Venepuncture was performed by a certified phlebotomist. Blood samples were collected in EDTA vacutainers, left at room temperature for 30 minutes, then centrifuged at 2000 g for 10 minutes at 4°C. The resulting plasma was aliquoted and stored at –80°C. Genomic DNA was extracted from whole blood samples, collected into EDTA vacutainers(Becton Dickinson) using the QIAamp DNA Blood Maxi Kit(Qiagen). The concentration and purity of the DNA were assessed using a Nanodrop One spectrophotometer (Thermo Fisher Scientific, United Kingdom). DNA was analysed using either the StepOne plus or QuantStudio 7 Pro Real-Time Polymerase Chain Reaction analyser(Applied Biosystems) to determine the allelic variants of APOE. APOE genotypes were determined using SNPs rs429358 and rs7412(Life Technologies), as per manufacturer’s protocol, using a 96-well MicroAmp Fast(StepOne Plus)/Optical(Quanstudio) reaction plate(Life Technologies), with 10ng of DNA and either one of the 2 SNPs, in a 10ul TaqPath ProAmp master mix reaction(Applied Biosystems)^63,64^.

Results were analysed using the Design and Analysis 2.5.1 Real Time PCR system software(Applied Biosystems) and respective APOE genotype were rated independently by raters. Quanterix’s Single Molecule Array(Simoa) digital biomarker technology platform was used to quantify all plasma biomarkers(NfL, GFAP, Aβ40, Aβ42, p-tau181)(Quanterix, Billerica, MA, USA). The Neurology 4- PlexE(NfL, GFAP, Aβ40, Aβ42) and p-tau181 Advantage V2.1 kits were utilized for expression analysis on the HD-X Analyzer platform(Quanterix, Billerica, MA, USA) in accordance with manufacturer’s protocol^65^.

### Statistical Analyses

Continuous variables were summarized using means and standard deviations, and categorical variables using frequencies and percentages. To address skewed distributions, blood biomarkers and behavioral data underwent logarithmic transformations, with a constant of 1 added to variables containing values below 1 to facilitate this transformation.

Comparisons between CN and MCI groups were performed using two independent sample t-test for continuous variables and Chi-square tests for categorical variables. Variance inflation factor (VIF) statistics were assessed to ensure no multicollinearity among predictor variables, with all included covariates demonstrating VIF values below 10.

Multivariable logistic regression analyses were executed using a backward variable selection approach in two separate models: Model 1 evaluated behavioral parameters (MBI-C, DASS), while Model 2 examined BBM (NfL, GFAP, Aβ40, Aβ42, p-tau181). A final comprehensive model (Model 3), using enter method, combined significant variables from Models 1 and 2 to ascertain those variables most strongly associated with MCI diagnosis, adjusting for age, years of education, gender, and APOE status. Results from logistic regression analyses were expressed as odds ratios (OR), accompanied by 95% confidence intervals (CI) and corresponding p-values. The discriminative capability of each logistic regression model (Models 1, 2, and 3) was further evaluated through receiver operating characteristic (ROC) curve analyses, with areas under the curve (AUC) computed. Model goodness-of-fit was evaluated using the Hosmer-Lemeshow test.

To further understand the association between behavioral parameters and cognitive diagnoses with BBM outcome, additional logistic regression analyses were implemented. Predictors were behavioural-biomarkers as continuous variables, adjusting for demographics, APOE status, and cognitive diagnosis (Model 4). Outcomes were blood biomarker status (i.e., high vs low), based on dichotomization using a median split as used in previous studies^66–68^.

Given the exploratory nature of these analyses, corrections for multiple comparisons were not applied. Results were interpreted based on effect sizes, CIs, and a significance criterion set at p < 0.05, aligned with standards adopted by previous exploratory studies^69^. All analyses were performed using IBM SPSS Statistics (Version 29.0, Armonk, NY: IBM Corp.). The sample size and study design of the BIOCIS study provided adequate statistical power for primary endpoints and supported the validity of these secondary exploratory analyses involving behavioral biomarkers, blood biomarkers, and cognitive outcomes.

## Results

### Demographics

Of the 1468 participants, 678 were classified as CN or as having MCI, and had complete data on NPS, blood-based biomarkers BBMs, and neuropsychological assessments. These participants were included in the final analysis. The inclusion process is outlined in Figure 1.

**Figure 1.**
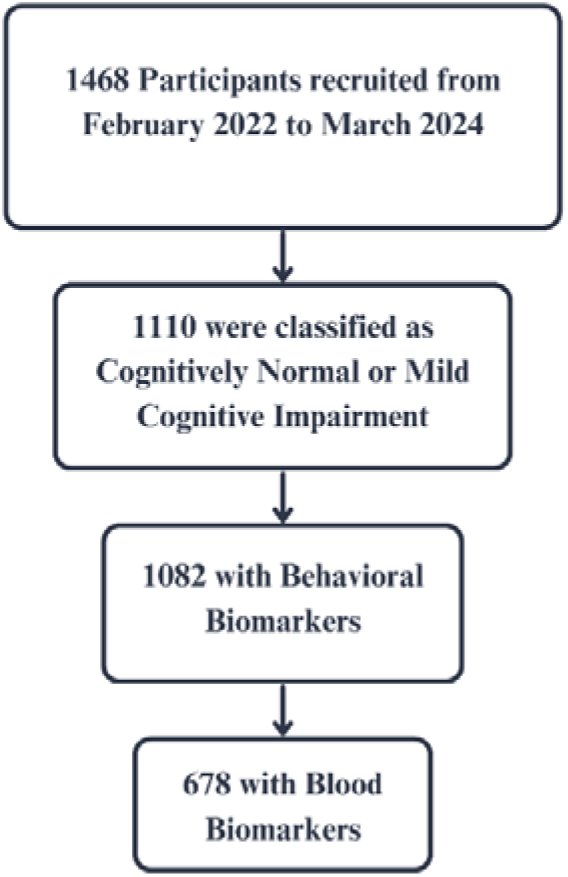
Flowchart of Inclusion Criteria for Participants

**Figure 2.**
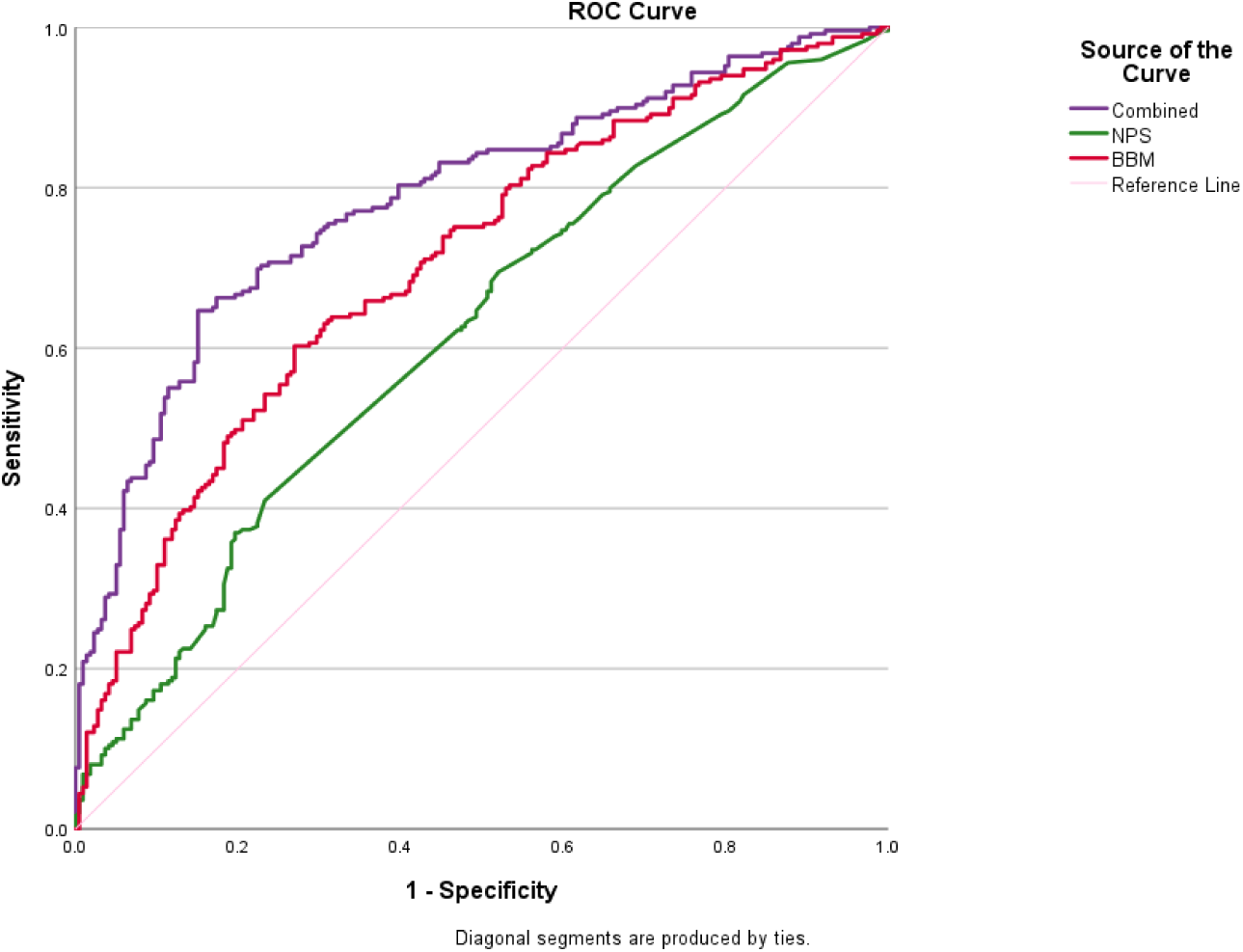
Receiver Operating Characteristic (ROC) Curves Comparing Different Models for Distinguishing Between Cognitively Normal and Mild Cognitive Impairment Groups. Abbreviations: BBM = Blood biomarkers, NPS = Neuropsychiatric symptoms

The MCI group was older (p<.001) and had fewer years of education (p<0.001). For behavioral assessments, MBI-C-Total scores were higher in the MCI group (p=0.006), with significant differences in the interest (p=0.005), mood (p=0.005), and control (p=0.009) subdomains.

Additionally, BBM analyses showed higher mean levels of Aβ40 (p< 0.001), NfL (p< 0.001), and GFAP (p< 0.001) in the MCI group compared to CN. Table 1 summarizes demographic, NPS and BBM data.

**Table 1.** Participant demographics, behavioral, and blood biomarker levels by cognitive diagnosis.

| Variable | Total Cohort<br>N=678 | Cognitively Normal<br>N=334 | Mild Cognitive<br>Impairment<br>N=344 | p-value* |
| --- | --- | --- | --- | --- |
| **Demographics** |  |  |  |  |
| Age (years) † | 59.16 ± 11.02 | 54.89 ± 10.24 | 63.31 ± 10.14 | < 0.001 |
| Gender (male) □ | 411 (60.50%) | 208 (62.00%) | 203 (59.00%) | 0.432 |
| Education (years) † | 14.76 ± 3.69 | 15.77 ± 3.33 | 13.78 ± 3.76 | < 0.001 |
| Ethnicity |  |  |  | 0.572 |
| Chinese | 593 (87.5%) | 256 (76.6%) | 337 (97.9%) |  |
| Malay | 18 (2.7%) | 10 (3.0%) | 8 (2.3%) |  |
| Indian | 39 (5.8%) | 18 (5.4%) | 21 (6.1%) |  |
| Others | 25 (3.7%) | 12 (3.6%) | 13 (3.8%) |  |
| Presence of APOE ε4 □ | 119 (17.70%) | 54 (16.20%) | 65 (19.00%) | 0.471 |
| **Behavioral biomarkers** |  |  |  |  |
| **Mild Behavioral Impairment-Checklist** |  |  |  |  |
| Total† | 2.18 ± 5.22 | 1.63 ± 3.66 | 2.73 ± 6.36 | 0.006 |
| Interest† | 0.42 ± 1.30 | 0.28 ± 0.86 | 0.56 ± 1.61 | 0.005 |
| Mood† | 0.60 ± 1.56 | 0.43 ± 1.26 | 0.76 ± 1.79 | 0.005 |
| Control† | 0.72 ± 1.64 | 0.55 ± 1.22 | 0.89 ± 1.95 | 0.009 |
| Social† | 0.20 ± 0.63 | 0.19 ± 0.58 | 0.21 ± 0.68 | 0.616 |
| Beliefs† | 0.24 ± 3.08 | 0.17 ± 2.19 | 0.30 ± 3.75 | 0.587 |
| **Depression Anxiety Stress Scale** |  |  |  |  |
| Depression†<sup>∇</sup> | 3.41 ± 4.63 | 2.92 ± 4.23 | 3.88 ± 4.94 | 0.007 |
| Anxiety†<sup>∇</sup> | 4.00 ± 4.19 | 3.78 ± 4.08 | 4.22 ± 4.30 | 0.174 |
| Stress†<sup>∇</sup> | 6.31 ± 5.92 | 6.21 ± 5.74 | 6.40 ± 6.10 | 0.671 |
| **Blood biomarkers** |  |  |  |  |
| Aβ42 (pg/mL)†<sup>∇</sup> | 5.22 ± 2.27 | 5.10 ± 1.91 | 5.34 ± 2.57 | 0.167 |
| Aβ40 (pg/mL)†<sup>∇</sup> | 78.76 ± 24.81 | 74.68 ± 23.28 | 82.73 ± 25.63 | < 0.001 |
| Aβ42/40 (ratio)†<sup>^</sup> | 0.0672 ±<br>0.0241 | 0.0687 ± 0.0201 | 0.0658 ± 0.0274 | 0.110 |
| p-tau181 (pg/mL)†<sup>∇</sup> | 18.67 ± 10.13 | 18.15 ± 11.86 | 19.16 ± 8.08 | 0.194 |
| p-tau181/Aβ42(ratio)†<sup>∇</sup> | 4.23 ± 3.13 | 4.19 ± 3.54 | 4.26 ± 2.69 | 0.780 |
| NfL (pg/mL)†<sup>^</sup> | 14.45 ± 8.73 | 12.18 ± 6.96 | 16.66 ± 9.67 | < 0.001 |
| GFAP (pg/mL)†<sup>^</sup> | 84.90 ± 44.38 | 71.41 ± 32.29 | 98.00 ± 50.26 | < 0.001 |

Note. † = mean ± Standard Deviation, □ = Frequency (percentages), ^ = higher values correspond to better scores, ∇ = lower values correspond to better scores Abbreviations: APOE = Apolipoprotein E, Aβ = Amyloid Peptides, p-tau181 = Phosphorylated Tau at position 181, GFAP = Glial fibrillary acidic protein, NfL = Neurofilament Light Chain

### Behavioral and Blood-Based Biomarkers Associated with MCI Compared to CN

Table 2 summarizes the results of the multivariable logistic regression analyses, detailing significant behavioral and blood-based variables associated with MCI compared to CN.

**Table 2.** Logistic Regression Models Identifying Behavioral and Blood Biomarkers associated with Mild Cognitive Impairment diagnosis.

| Variable | Odds Ratio | 95% CI | p-value |
| --- | --- | --- | --- |
| **Model 1 – Behavioral Biomarkers, AUC = 0.593** |  |  |  |
| MBI-C-Mood | 1.587 | (1.127, 2.236) | **0.008** |
| DASS-Depression | 1.250 | (1.013, 1.542) | **0.037** |
| DASS-Stress | 0.800 | (0.660, 0.970) | **0.023** |
| **Model 2 – Blood Biomarkers, AUC = 0.697** |  |  |  |
| NfL | 2.096 | (1.365, 3.217) | **0.001** |
| GFAP | 2.987 | (1.913, 4.664) | **<0.001** |
| **Model 3 – Combining Behavioral and Blood Biomarkers, AUC = 0.786** |  |  |  |
| GFAP | 3.636 | (1.959, 6.751) | **<0.001** |
| MBI-C-Mood | 2.614 | (1.538, 4.441) | **< 0.001** |

*Note.* Outcome = Mild Cognitive Impairment (reference = cognitively normal). Only predictors retained after backward elimination are shown. Backward stepwise selection; Enter method.
Model 1 initial predictors: MBI-C subdomains (Interest, Mood, Control, Social, Beliefs) and DASS sub-scales (Depression, Anxiety, Stress).
Model 2 initial predictors: NfL, GFAP, Aβ 40, Aβ 42, p-tau181.
Abbreviations: Aβ = Amyloid Peptides, AUC = Area under the curve, DASS = Depression Anxiety Stress Scales, GFAP = Glial fibrillary acidic protein, MBI-C = Mild Behavioral Impairment-Checklist, NfL = Neurofilament Light Chain, p-tau181 = Phosphorylated Tau at position 181

In the behavioral model (Model 1), for each 1-point increase in the MBI-C-Mood subdomain, the odds of being MCI over being CN increased by approximately 59% (OR: 1.587, 95% CI: 1.127– 2.236, p=0.008). Similarly, for each 1-point increase in the DASS Depression score, the odds of MCI over CN increased by 25% (OR: 1.250, 95% CI: 1.013–1.542, p=0.037). In contrast, higher DASS-Stress scores were inversely associated with cognitive impairment, with each 1-point increase in DASS-Stress reducing the odds of MCI by 20% (OR: 0.800, 95% CI: 0.660–0.970, p=0.023). The AUC for behavioral-markers alone was 0.593 (95% CI: 0.55–0.636).

Among BBM (Model 2), elevated NfL and GFAP levels were strongly associated with the likelihood of being classified as MCI over CN. Each unit increase in NfL corresponded to a 110% increase in MCI likelihood over CN (OR: 2.096, 95% CI: 1.365–3.217, p=0.001). GFAP showed an even stronger association, with each unit increase raising the odds of MCI over CN by approximately 199% (OR: 2.987, 95% CI: 1.913–4.664, p<0.001).

### Combined Model of Predictive Value of Behavioral Scores and Blood Biomarker Levels for Cognitive Status

In the combined behavioral and blood model (Model 3), variables from Table 2(i.e., variables from the Model 1 and 2 meeting statistical significance) were included along with demographics and APOE status as clinical confounders. Higher GFAP (OR: 3.636, 95% CI: 1.959–6.751, p< 0.001) and MBI- C-Mood(OR: 2.614, 95% CI: 1.538–4.441, p< 0.001) significantly increased the likelihood of MCI by 263.6% and 161.4% respectively. The combined Model 3’s AUC was 0.786 (95% CI: 0.744–0.827), surpassing separate models based on either Model 1 (NPS; AUC: 0.593, 95% CI: 0.55–0.636) or Model 2 (BBM; AUC: 0.697, 95% CI: 0.658–0.736).

### Behavioral Markers in Association with Dichotomized BBM Levels

For Model 4, with continuous behavior scores as predictors and biomarker status as a binary outcome, a higher MBI-C-Total score was significantly associated with odds of high Aβ42 status (OR: 1.437, 95% CI: 1.127–1.831, p=0.003). Specific MBI-C subdomains were significantly linked to likelihood of high Aβ42: MBI-C-Interest(OR: 1.800, 95% CI: 1.139–2.845, p=0.012), MBI-C-Mood (OR: 1.587, 95% CI: 1.068–2.359, p=0.022), and MBI-C-Control (OR: 1.782, 95% CI: 1.242–2.557, p=0.002).

Higher DASS-Stress scores were significantly associated with likelihood of high Aβ40 levels (OR: 1.252, 95% C I: 1.039–1.508, p=0.018). MBI-C-Total score was associated with likelihood of high p- tau181 (OR: 1.279, 95% CI: 1.001–1.634, p=0.049). Additionally, higher DASS-Depression scores(OR: 1.302, 95% CI: 1.061–1.598, p=0.012) and higher DASS-Stress scores (OR: 1.238, 95% CI: 1.020–1.503, p=0.031) significantly increased the odds of high p-tau181. Lastly, higher DASS-Anxiety scores were marginally associated with probability of high GFAP levels(OR: 1.260, 95% CI: 1.000–1.587, p=0.05). Table 3 presents a detailed summary of results.

**Table 3.** Multivariable Logistic regression associations between Behavioral measures and dichotomized (median-split) Blood biomarkers.

| Variable | Aβ42<sup>†</sup> | Aβ40<sup>†</sup> | Aβ42/40<sup>†</sup> | p-tau181<sup>†</sup> | p-tau181/Aβ42<sup>†</sup> | Nf-L<sup>†</sup> | GFAP<sup>†</sup> |
| --- | --- | --- | --- | --- | --- | --- | --- |
| **MBI-C Total** | **1.44 (1.13, 1.83)\*\*** | 1.16 (0.92, 1.47) | 1.15 (0.86, 1.53) | **1.28 (1.001, 1.63)\*** | 0.84 (0.66, 1.07) | 0.83 (0.62, 1.12)<sup>†</sup> | 1.08 (0.83, 1.39) |
| **MBI-C Interest** | **1.8 (1.14, 2.85)\*** | 1.34 (0.87, 2.06) | 1.21 (0.72, 2.02) | 1.34 (0.86, 2.07) | 0.78 (0.51, 1.20) | 0.69 (0.40, 1.20) | 1.16 (0.74, 1.83) |
| **MBI-C Mood** | **1.59 (1.07, 2.36)\*** | 1.21 (0.82, 1.77) | 1.43 (0.87, 2.34) | 1.27 (0.85, 1.89) | 0.82 (0.55, 1.20) | 0.84 (0.52, 1.37) | 1.32 (0.87, 1.99) |
| **MBI-C Control** | **1.78 (1.24, 2.56)\*\*** | 1.34 (0.95, 1.88) | 1.11 (0.74, 1.67) | 1.37 (0.97, 1.94) | 0.79 (0.56, 1.12) | 0.84 (0.55, 1.28) | 1.05 (0.73, 1.52) |
| **MBI-C Social** | 1.45 (0.68, 3.09) | 1.37 (0.64, 2.90) | 1.16 (0.46, 2.92) | 1.45 (0.66, 3.16) | 0.93 (0.44, 1.99) | 0.87 (0.34, 2.25) | 0.90 (0.40, 2.01) |
| **MBI-C Beliefs** | 0.68 (0.20, 2.26) | 0.76 (0.22, 2.64) | 3.01 (0.38, 23.6) | 3.34 (0.91, 12.3) | 1.26 (0.37, 4.35) | 0.46 (0.08, 2.52) | 0.97 (0.26, 3.67) |
| **DASS Depression** | 1.09 (0.90, 1.32) | 1.22 (<1, 1.48) | 1.18 (0.94, 1.50) | **1.30 (1.06, 1.60)\*** | 0.97 (0.80, 1.18) | 0.83 (0.65, 1.06) | 1.09 (0.88, 1.35) |
| **DASS Anxiety** | 1.17 (0.95, 1.44) | 1.23 (<1, 1.52) | 0.98 (0.76, 1.25) | 1.13 (0.91, 1.41) | 0.95 (0.77, 1.17) | 0.93 (0.72, 1.20) | 1.26 (<1, 1.59) |
| **DASS Stress** | 1.20 (1.004, 1.45)* | **1.25 (1.04, 1.51)\*** | 1.01 (0.81, 1.26) | **1.24 (1.02, 1.50)\*** | 0.98 (0.81, 1.18) | 1.07 (0.86, 1.34) | 1.04 (0.85, 1.27) |

Note. <sup>†</sup>Odds ratio (95% CI), \* p<0.05, \*\* p<0.005. Significant associations are bolded. Odds ratios represent the change in odds of being above the median for each blood biomarker per unit increase in the behavioral score. All models were adjusted for age, years of education, gender, APOE ε4 carrier status, and cognitive diagnosis.
Abbreviations: Aβ =Amyloid peptides; DASS=Depression Anxiety Stress Scales; GFAP=Glial fibrillary acidic protein; MBI-C=Mild Behavioural Impairment Checklist; NfL=Neurofilament light chain; p-tau181=Phosphorylated tau 181.

## Discussion

### Integration of Biomarkers for Early Detection of Cognitive Impairment

Our findings demonstrate that combining plasma biomarkers with neuropsychiatric assessments significantly improves the identification of MCI, compared to using either modality in isolation. The comparison of models showed that the integrated model (NPS + BBM) had the highest accuracy (AUC ∼0.79), outperforming models based solely on BBM (AUC ∼0.70) or solely on NPS (AUC ∼0.59). Three key insights emerge from these results: (1) Specific behavioral changes, particularly mood-related symptoms, are strongly linked to cognitive impairment; (2) Blood biomarkers, especially those reflecting neuroinflammation, are associated with MCI status; and (3) Critically, combining NPS and BBM yields superior predictive power, highlighting that each modality offers unique and complementary information for the early detection of cognitive decline.

### NPS as Early Indicators of MCI

Higher MBI-C total scores, particularly in the Mood, Interest, and Control subdomains, were associated with increased likelihood of MCI. The strong predictive association between mood disturbances and cognitive impairment is warrants attention. Mood symptoms often precede overt cognitive decline and likely reflect dysfunction in frontal-limbic circuitry, including regions such as the orbitofrontal cortex, amygdala, and hippocampus, known to be affected early in Alzheimer’s pathology^70,71^. Notably, in our study MBI-C Mood had greater association strength with MCI diagnosis than the general DASS Depression scale, likely because MBI-C specifically captures persistent, late-onset mood changes rather than transient depressive symptoms^43^.

In addition to mood symptoms, apathy and impulse dyscontrol were elevated in individuals with MCI in univariate analyses, although these associations did not persist in the multivariate model. Still, these findings are clinically meaningful. Apathy is among the earliest and most prevalent NPS in prodromal AD, linked consistently to dysfunction in fronto-striatal networks and posterior cingulate cortex hypometabolism^72^. Similarly, impulse dyscontrol—including irritability, agitation, and aberrant reward-seeking behaviors—may indicate early frontal disinhibition or subtle executive dysfunction, commonly occurring with frontal-subcortical network impairment secondary to AD or vascular pathology^73^. Collectively, the association between these behavioral changes and cognitive deficits reinforces the conceptualization of MBI as a prodromal phase within the Alzheimer’s disease continuum.

Interestingly, we found an inverse relationship between self-reported stress (DASS-Stress) and the likelihood of MCI. Two non mutually exclusive mechanisms may explain this counter intuitive pattern. First, prodromal cognitive decline can blunt emotional reactivity and reduce insight, leading individuals with incipient neurodegeneration to under report stress related affect. This interpretation is consistent with reports of attenuated emotional responsivity in early AD^74^. Second, it is possible that cognitively intact adults remain more professionally and socially active, which may expose them to a greater number of daily stressors. In contrast, individuals with MCI may be more likely to retire or socially withdraw, resulting in fewer stress-inducing experiences. Together, these observations highlight the symptom-specific and potentially bidirectional nature of neuropsychiatric– cognitive relationships, underscoring the importance of interpreting behavioural scores within the broader context of an individual’s cognitive and functional status.

Taken together, our findings underscore behavioral changes—particularly mood disturbances—as valuable indicators of incipient cognitive impairment. Current research supports this view—NPS are not merely reactive to cognitive decline, but may precede and actively contribute to its progression ^33,35,43,46^. Chronic affective symptoms can induce hypothalamic-pituitary-adrenal(HPA)-axis dysregulation, elevate cortisol levels, and trigger neuroinflammatory processes, all of which exacerbate Alzheimer’s pathology and neuronal injury^75–77^. Thus, NPS could reflect underlying neurodegenerative processes while simultaneously accelerating their progression, creating a detrimental feedback loop.

### Behavioral-Biomarkers as Early Indicators of Neurodegeneration

Higher MBI-C scores were significantly associated with elevated Aβ42 levels, particularly within the subdomains of Interest, Mood, and Control, supporting the role of early behavioral disturbances as clinical indicators of amyloid-related pathology^43,58,78,79^. Moreover, higher DASS-Stress scores were strongly associated with elevated Aβ40 levels, reinforcing evidence of an association between stress and amyloid pathology^80,81^. The inconsistent findings for Aβ42 and the Aβ42/Aβ40 ratio in our study and throughout literature highlight variability likely attributable to differences in study cohorts and methodological heterogeneity, underscoring the complexity of utilizing plasma amyloid biomarkers in dementia research.

Beyond amyloid markers, elevated MBI-C Total scores, DASS-Depression and DASS-Stress scores were significantly linked to higher p-tau181 levels, aligning with emerging evidence that behavioral symptoms may correlate with tau pathology^43–45,82,83^. This relationship illustrates the potential interplay between stress, depression, and tau-related neurodegeneration, which is critical for understanding the broader neuropathological mechanisms underlying cognitive decline. Finally, a marginal association between elevated GFAP levels and DASS-Anxiety scores suggests a tentative link between anxiety and neuroinflammatory processes, potentially reflecting glial activation in response to chronic psychological distress.

### Plasma Biomarkers and MCI status

Our results demonstrate the potential of BBM to distinguish MCI from CN participants, supporting their utility in early detection of AD and related cognitive disorders.

### Amyloid Pathology and Early Detection

Although the Aβ42/40 ratio did not significantly differ between the CN and MCI groups, the elevated Aβ40 levels in MCI participants suggest early amyloid dysregulation. Aβ40, which is increasingly implicated in amyloid plaque formation and neurotoxicity, may serve as an early biomarker of amyloid pathology despite historically receiving less attention than Aβ42^84^. This pattern aligns with recent research indicating that Aβ40 elevations may precede Aβ42 changes^85^, affirming their role as an early biomarker in NDD. Notably, plasma amyloid markers were not retained in the final combined model, suggesting a weaker association with cognitive impairment relative to other BMM. This finding may reflect the lower prevalence of APOE ε4 carriers in our cohort.^60^.

### Tau

P-tau181 did not differ significantly between cognitive groups, nor predict MCI status. Although tau pathology is central to AD, plasma p-tau181 may have lower sensitivity to early cognitive impairment compared to markers of amyloid dysregulation or neuroinflammation^86^. This corresponds with recent findings that p-tau181 becomes more relevant in later AD stages as tau pathology accumulates in the medial temporal cortex and spreads to other cortical regions^87,88^. Thus, the limited discriminative ability of plasma p-tau181 in our study may reflect the preclinical or prodromal disease stage of participants, where tau-related changes are still emerging and less detectable in peripheral measures.

### Neurodegeneration

MCI participants had significantly higher plasma NfL levels, reinforcing NfL’s role as a sensitive marker of axonal injury and early neurodegeneration ^89–91^. Elevated plasma NfL has consistently predicted progression from MCI to dementia across diverse populations ^92^. Our findings further support NfL as a robust indicator of underlying disease processes, helping to identify individuals on the neurodegenerative trajectory.

### Neuroinflammation

In our study, GFAP emerged as the most robust blood-based biomarker associated with cognitive impairment, underscoring the involvement of astrocytic activation in early neurodegenerative processes. As a well-established marker of reactive astrogliosis, GFAP reflects astrocyte responses to amyloid accumulation and other neuropathological insults^93^. Mechanistically, reactive astrocytes contribute to neuroinflammation by releasing pro-inflammatory cytokines and glutamate, leading to synaptic dysfunction and neuronal injury^94,95^. Correspondingly, we observed significantly elevated GFAP levels in individuals with MCI compared to cognitively normal participants, supporting its role in prodromal disease stages. This elevation may be driven in part by the higher cerebrovascular disease burden in our population^60,96^, which contributes and amplifies neuroinflammation^97,98^.

Our BBM findings confirm that plasma markers, particularly GFAP, have strong potential to detect the biological underpinnings of MCI. However, while BBMs provide valuable insights into neurodegenerative pathology, they do not fully capture the clinical manifestations or subjective experiences of individuals. Since early cognitive decline often presents subtly at the behavioral level, combining NPS assessments with BBMs offers a more comprehensive and clinically relevant framework for identifying and managing prodromal cognitive impairment.

### Enhanced Predictive Power through Integrated Models

The most novel aspect of this study is demonstrating the added value of a multimodal approach that integrates BBM with NPS assessment. Previous work in AD has shown that combining cognitive test scores with BBM yields higher accuracy for detecting AD dementia than either alone^99,100^. We extend this principle by demonstrating that combining NPS with BBM improves early identification of cognitive impairment secondary to NDD, with their convergence offering the strongest distinction between MCI and cognitively normal individuals. BBM and NPS capture complementary aspects of disease progression: BBM contributes biological specificity, while NPS enhance clinical sensitivity. BBM detect underlying AD and NDD pathology, but some individuals remain asymptomatic despite abnormal biomarkers^101^. In contrast, NPS reflect structural and functional changes in brain circuits regulating behavior and emotion, are associated with structural indicators of neurodegeneration, and have been shown to precede the emergence of cognitive symptoms^43,77,102,103^.

### The Role of Neuroinflammation and Behavioral Changes as Correlates of Cognitive Impairment

The prominent role of GFAP in our findings emphasizes neuroinflammation as a key process in early pathogenesis of NDD. interventions aimed at modulating astrocytic responses prior to irreversible neuronal damage. Elevated GFAP reflects astrocytic activation that not only signals neuronal stress but also promotes progression through cytokine release and reactive gliosis, exacerbating synaptic injury and cognitive decline^104,105^. Consequently, GFAP may serve as a vascular-related marker of astrocytic stress and delimit a therapeutic window for anti-inflammatory intervention before irreversible damage ensues.

The significant predictive value of MBI-C Mood scores highlights mood disturbances as potential early markers of neurodegeneration. Rather than reflecting downstream psychological effects of cognitive decline, emotional dysregulation symptoms may signal distinct biological processes beyond core AD pathology, as recently highlighted by Rabl *et al.*^48^. Converging evidence links elevated mood disturbances and plasma GFAP elevations with neurodegenerative markers, particularly amyloid pathology and cortical atrophy^30,68,106,107^, suggesting overlapping biological substrates.

In the combined model, both GFAP and MBI-C Mood remained significant, suggesting an interplay between neuroinflammation and affective symptoms in early stages of cognitive impairment.

Neuroinflammatory changes—potentially triggered by amyloid deposition or other insults—may contribute to mood disturbances by disrupting neurotransmitter systems and promoting cytokine-mediated alterations in neuronal function and connectivity^108^, and growing evidence support associations between affective symptoms and neuroinflammatory markers. Our findings highlight elevated GFAP and mood dysregulation as interrelated and potentially modifiable therapeutic targets. Cognitive behavioural therapy (CBT) has demonstrated efficacy in alleviating depressive symptoms in individuals with dementia ^109^. While no current therapies directly target plasma GFAP or astrocytic activation, emerging approaches—such as amyloid-lowering agents like lecanemab—may indirectly attenuate neuroinflammation^110^. Prospective, mechanism focused trials are required to determine whether dual targeted strategies confer additive protection beyond either approach alone.

### Limitations

As with most studies, several limitations must be considered. The exclusion of participants with severe psychiatric conditions may limit the generalizability of our findings, as this group could offer valuable insights into early cognitive and behavioral changes. The cross-sectional design restricts our ability to infer causality or to track longitudinal changes in biomarkers and cognitive outcomes. Furthermore, other important confounding factors, including cardiometabolic conditions, brain imaging measures, and lifestyle factors, were not accounted for, which might influence biomarker levels and cognitive performance. The absence of PET or CSF data precludes direct correlation between BBM and established neurodegeneration markers, limiting biological validation of our findings. The relatively small sample size also reduces statistical power, limiting generalizability to broader and more diverse populations. Multiple comparison corrections were not applied, warranting cautious interpretation and independent replication to minimize false-positive results. Future longitudinal studies, such as those planned within the expanded BIOCIS cohort, are essential to validate these associations and elucidate temporal relationships among NPS, BBM, and cognitive impairment. Finally, translating these findings into clinical practice will require research focused on confirming biomarker specificity and mechanisms, evaluating predictive value for disease progression, and ensuring scalability, cost-effectiveness, and accessibility of biomarker assays for implementation in global healthcare settings.

## Conclusions

In summary, this study supports the integration of biological and behavioural markers for a novel approach in early detection of NDD. In our multi-ethnic cohort, elevated plasma GFAP and higher MBI-C Mood scores were the strongest joint indicators of MCI, underscoring the intertwined roles of neuroinflammation and affective changes. An integrative approach offers translational pathways - early stratification (based on BMM) enables enrollment in disease-modifying trials, while targeted interventions for poor mood and emotional dysregulation may confer additional cognitive benefit.

Stratifying patients by combined biomarker and clinical profiles aligns with precision medicine goals and lays the groundwork for individualized monitoring and therapeutic planning. As the field shifts toward earlier intervention, such models will be crucial for translating pathophysiological findings into clinical practice.

## Data Availability

Data will be made available upon reasonable request to the corresponding author.

## Abbreviations

AD: Alzheimer’s Disease
ADNI: Alzheimer’s Disease Neuroimaging Initiative
Aβ40: Amyloid-Beta 40
Aβ42: Amyloid-Beta 42
APOE ε4: Apolipoprotein E ε4
ATN: Amyloid, Tau, and Neurodegeneration
AUC: Area Under the Curve
BBM: Blood-Based Biomarkers
BIOCIS: Biomarkers and Cognition Study, Singapore
CN: Cognitively Normal
CSF: Cerebrospinal Fluid
DASS: Depression, Anxiety, and Stress Scales
EDTA: Ethylenediaminetetraacetic Acid
GFAP: Glial Fibrillary Acidic Protein
MBI: Mild Behavioral Impairment
MBI-C: Mild Behavioral Impairment Checklist
MCI: Mild Cognitive Impairment
NDD: Neurodegenerative Disease
NfL: Neurofilament Light
NIA-AA: National Institute on Aging-Alzheimer’s Association
NPS: Neuropsychiatric Symptoms
PCR: Polymerase Chain Reaction
PET: Positron Emission Tomography
p-tau181: Phosphorylated Tau at position 181
ROC: Receiver Operating Characteristic
Simoa: Single Molecule Array
SNPs: Single Nucleotide Polymorphisms

## Declarations Ethics approval and consent to participate

Informed consent was obtained from all participants according to the Declaration of Helsinki and local clinical research regulations, and procedures used in the study were in accordance with ethical guidelines. This study has been approved by the NTU Institutional Review Board (NTU-IRB-2021-1036), and procedures used in the study were in accordance with ethical guidelines.

## Consent for publication

Not applicable.

## Availability of data and materials

The datasets are available from the corresponding author on reasonable request.

## Acknowledgements

The authors express their gratitude to all individuals who are currently or will be participating in research at the Dementia Research Centre (Singapore).

## Author Contributions

### Concept and design

Yi Jin Leow

### Acquisition, analysis, or interpretation of data

Yi Jin Leow, Zahinoor Ismail, Seyed Ehsan Saffari, Gurveen Kaur Sandhu, Pricilia Tanoto, Faith Phemie Hui En Lee, Smriti Ghildiyal, Shan Yao Liew, Gursimar Bhalla, Sim Xin Ying, Adnan Azam Mohammed, Ashwati Vipin, Chao Dang, Nagaendran Kandiah

### Drafting of the manuscript

Yi Jin Leow, Zahinoor Ismail, Seyed Ehsan Saffari, Nagaendran Kandiah

### Statistical analysis

Yi Jin Leow, Seyed Ehsan Saffari

### Supervision

Nagaendran Kandiah

## References

1. Klyucherev TO, Olszewski P, Shalimova AA, et al. Advances in the development of new biomarkers for Alzheimer’s disease. Translational neurodegeneration. 2022;11(1):25.

2. Li B, He Y, Ma J, et al. Mild cognitive impairment has similar alterations as Alzheimer’s disease in gut microbiota. Alzheimer’s & Dementia. 2019;15(10):1357–1366.

3. Aye S, Handels R, Winblad B, Jönsson L. Optimising Alzheimer’s disease diagnosis and treatment: assessing cost-utility of integrating blood biomarkers in clinical practice for disease-modifying treatment. The Journal of Prevention of Alzheimer’s Disease. Published online 2024:1–15.

4. Blennow K, Galasko D, Perneczky R, et al. The potential clinical value of plasma biomarkers in Alzheimer’s disease. Alzheimer’s & Dementia. 2023;19(12):5805–5816.

5. Dubois B, Villain N, Schneider L, et al. Alzheimer disease as a clinical-biological construct—an International Working Group recommendation. JAMA neurology. Published online 2024.

6. Hansson O, Edelmayer RM, Boxer AL, et al. The Alzheimer’s Association appropriate use recommendations for blood biomarkers in Alzheimer’s disease. Alzheimer’s & Dementia. 2022;18(12):2669–2686.

7. Kodosaki E, Zetterberg H, Heslegrave A. Validating blood tests as a possible routine diagnostic assay of Alzheimer’s disease. Expert Review of Molecular Diagnostics. 2023;23(12):1153–1165.

8. Kivisäkk P, Carlyle BC, Sweeney T, et al. Plasma biomarkers for diagnosis of Alzheimer’s disease and prediction of cognitive decline in individuals with mild cognitive impairment. Frontiers in neurology. 2023;14:1069411.

9. Simrén J, Leuzy A, Karikari TK, et al. The diagnostic and prognostic capabilities of plasma biomarkers in Alzheimer’s disease. Alzheimer’s & Dementia. 2021;17(7):1145–1156.

10. Palmqvist S, Tideman P, Mattsson-Carlgren N, et al. Blood biomarkers to detect Alzheimer disease in primary care and secondary care. JAMA. 2024;332(15):1245–1257.

11. 11. CN Diagnostics. CN Diagnostics releases the PrecivityAD2^TM^ blood test for clinical care, a robust assay with high concordance to amyloid PET and CSF. August 17, 2024. Accessed December 4, 2025. https://c2n.com/news-releases/cnnbspdiagnostics-releases-the-precivityad2-blood-test-for-clinical-care

12. Quest Diagnostics. Quest Diagnostics launches new AD-Detect^TM^ blood test to aid in confirming Alzheimer’s disease. January 15, 2025. Accessed December 4, 2025. https://ir.questdiagnostics.com/press-releases/press-release-details/2025/Quest-Diagnostics-Launches-New-AD-Detect-Blood-Test-to-Aid-in-Confirming-Alzheimers-Disease/default.aspx

13. VandeVrede L, Rabinovici GD. Blood-Based Biomarkers for Alzheimer Disease—Ready for Primary Care? JAMA neurology. 2024;81(10):1030–1031.

14. Hansson O, Blennow K, Zetterberg H, Dage J. Blood biomarkers for Alzheimer’s disease in clinical practice and trials. Nature aging. 2023;3(5):506–519.

15. Li Y, Schindler SE, Bollinger JG, et al. Validation of plasma amyloid-β 42/40 for detecting Alzheimer disease amyloid plaques. Neurology. 2022;98(7):e688–e699.

16. Ankeny SE, Bacci JR, Decourt B, Sabbagh MN, Mielke MM. Navigating the Landscape of Plasma Biomarkers in Alzheimer’s Disease: Focus on Past, Present, and Future Clinical Applications. Neurology and Therapy. Published online 2024:1–17.

17. Cai H, Pang Y, Fu X, Ren Z, Jia L. Plasma biomarkers predict Alzheimer’s disease before clinical onset in Chinese cohorts. Nature Communications. 2023;14(1):6747.

18. Chen S, Huang Y, Shen X, Guo Y, Tan L, Dong Q. Longitudinal plasma phosphorylated tau 181 tracks disease progression in Alzheimer’s disease. Transl Psychiatry. 2021; 11: 356. Published online 2021.

19. Lu J, Ma X, Zhang H, et al. Head-to-head comparison of plasma and PET imaging ATN markers in subjects with cognitive complaints. Translational Neurodegeneration. 2023;12(1):34. doi:10.1186/s40035-023-00365-x

20. Chatterjee P, Pedrini S, Stoops E, et al. Plasma glial fibrillary acidic protein is elevated in cognitively normal older adults at risk of Alzheimer’s disease. Translational psychiatry. 2021;11(1):27.

21. Matthews DC, Kinney JW, Ritter A, et al. Relationships between plasma biomarkers, tau PET, FDG PET, and volumetric MRI in mild to moderate Alzheimer’s disease patients. Alzheimer’s & Dementia: Translational Research & Clinical Interventions. 2024;10(3):e12490.

22. Stevenson-Hoare J, Heslegrave A, Leonenko G, et al. Plasma biomarkers and genetics in the diagnosis and prediction of Alzheimer’s disease. Brain. 2023;146(2):690–699.

23. Zhu N, Santos-Santos M, Illán-Gala I, et al. Plasma glial fibrillary acidic protein and neurofilament light chain for the diagnostic and prognostic evaluation of frontotemporal dementia. Translational Neurodegeneration. 2021;10(1):50. doi:10.1186/s40035-021-00275-w

24. Chong JR, Hilal S, Tan BY, et al. Clinical utility of plasma p-tau217 in identifying abnormal brain amyloid burden in an Asian cohort with high prevalence of concomitant cerebrovascular disease. Alzheimer’s & Dementia. Published online 2025:e14502.

25. Liu E, Zhang Y, Wang JZ. Updates in Alzheimer’s disease: from basic research to diagnosis and therapies. Translational Neurodegeneration. 2024;13(1):45. doi:10.1186/s40035-024-00432-x

26. Crean S, Ward A, Mercaldi CJ, et al. Apolipoprotein E ε4 prevalence in Alzheimer’s disease patients varies across global populations: a systematic literature review and meta-analysis. Dementia and geriatric cognitive disorders. 2011;31(1):20–30.

27. Vipin A, Kumar D, Soo SA, et al. APOE4 carrier status determines association between white matter disease and grey matter atrophy in early-stage dementia. Alzheimer’s Research & Therapy. 2023;15(1):103.

28. Belloy ME, Andrews SJ, Le Guen Y, et al. APOE genotype and Alzheimer disease risk across age, sex, and population ancestry. JAMA neurology. 2023;80(12):1284–1294.

29. 29. EMA. Leqembi recommended for treatment of early Alzheimer’s disease. November 22, 2024. https://www.ema.europa.eu/en/news/leqembi-recommended-treatment-early-alzheimers-disease

30. Ng KP, Pascoal TA, Mathotaarachchi S, et al. Neuropsychiatric symptoms are early indicators of an upcoming metabolic decline in Alzheimer’s disease. Translational Neurodegeneration. 2021;10:1–11.

31. Paris A, Amirthalingam G, Karania T, et al. Depression and dementia: interrogating the causality of the relationship. *Journal of Neurology*, Neurosurgery & Psychiatry. Published online 2025.

32. Mortby ME, Burns R, Eramudugolla R, Ismail Z, Anstey KJ. Neuropsychiatric Symptoms and Cognitive Impairment: Understanding the Importance of Co-Morbid Symptoms. Journal of Alzheimer’s Disease. 2017;59(1):141–153. doi:10.3233/JAD-170050

33. Creese B, Brooker H, Ismail Z, et al. Mild behavioral impairment as a marker of cognitive decline in cognitively normal older adults. The American Journal of Geriatric Psychiatry. 2019;27(8):823–834.

34. Ismail Z, McGirr A, Gill S, Hu S, Forkert ND, Smith EE. Mild behavioral impairment and subjective cognitive decline predict cognitive and functional decline. Journal of Alzheimer’s disease. 2021;80(1):459–469.

35. McGirr A, Nathan S, Ghahremani M, Gill S, Smith EE, Ismail Z. Progression to dementia or reversion to normal cognition in mild cognitive impairment as a function of late-onset neuropsychiatric symptoms. Neurology. 2022;98(21):e2132–e2139.

36. Ismail Z, Agüera-Ortiz L, Brodaty H, et al. The Mild Behavioral Impairment Checklist (MBI-C): a rating scale for neuropsychiatric symptoms in pre-dementia populations. Journal of Alzheimer’s disease. 2017;56(3):929–938.

37. Ismail Z, Smith EE, Geda Y, et al. Neuropsychiatric symptoms as early manifestations of emergent dementia: provisional diagnostic criteria for mild behavioral impairment. Alzheimer’s & Dementia. 2016;12(2):195–202.

38. Leow Y, Soo SA, Kumar D, et al. Mild Behavioral Impairment and Cerebrovascular Profiles Are Associated with Early Cognitive Impairment in a Community-Based Southeast Asian Cohort. Journal of Alzheimer’s Disease. 2024;(Preprint):1–9.

39. Matsuoka T, Ismail Z, Narumoto J. Prevalence of mild behavioral impairment and risk of dementia in a psychiatric outpatient clinic. Journal of Alzheimer’s Disease. 2019;70(2):505–513.

40. Lovibond PF, Lovibond SH. The structure of negative emotional states: Comparison of the Depression Anxiety Stress Scales (DASS) with the Beck Depression and Anxiety Inventories. Behaviour research and therapy. 1995;33(3):335–343.

41. Freire ACC, Pondé MP, Liu A, Caron J. Anxiety and depression as longitudinal predictors of mild cognitive impairment in older adults. The Canadian Journal of Psychiatry. 2017;62(5):343–350.

42. Perin S, Lai J, Pase M, et al. Elucidating the association between depression, anxiety, and cognition in middle-aged adults: Application of dimensional and categorical approaches. Journal of Affective Disorders. 2022;296:559–566.

43. Ismail Z, Leon R, Creese B, Ballard C, Robert P, Smith EE. Optimizing detection of Alzheimer’s disease in mild cognitive impairment: a 4-year biomarker study of mild behavioral impairment in ADNI and MEMENTO. Molecular Neurodegeneration. 2023;18(1):50.

44. Ghahremani M, Wang M, Chen HY, et al. Plasma phosphorylated tau at threonine 181 and neuropsychiatric symptoms in preclinical and prodromal Alzheimer disease. Neurology. 2023;100(7):e683–e693.

45. Gonzalez Bautista E, Momméja M, de Mauléon A, et al. Mild behavioral impairment domains are longitudinally associated with pTAU and metabolic biomarkers in dementia free older adults. Alzheimer’s & Dementia. Published online 2024.

46. Creese B, Ismail Z. Mild behavioral impairment: measurement and clinical correlates of a novel marker of preclinical Alzheimer’s disease. Alzheimer’s research & therapy. 2022;14:1–5.

47. Gates N, Valenzuela M, Sachdev PS, Fiatarone Singh MA. Psychological well-being in individuals with mild cognitive impairment. Clinical interventions in aging. Published online 2014:779–792.

48. Rabl M, Clark C, Dayon L, Popp J. Neuropsychiatric symptoms in cognitive decline and Alzheimer’s disease: biomarker discovery using plasma proteomics. *Journal of Neurology*, Neurosurgery & Psychiatry. 2025;96(4):370–382.

49. Qiu WQ, Zhu H, Dean M, et al. Amyloid associated depression and ApoE4 allele: longitudinal follow up for the development of Alzheimer’s disease. International journal of geriatric psychiatry. 2016;31(3):316–322.

50. Pomara N, Bruno D, Plaska CR, et al. Plasma Amyloid-β dynamics in late-life major depression: a longitudinal study. Translational Psychiatry. 2022;12(1):301.

51. Blasko I, Kemmler G, Jungwirth S, et al. Plasma amyloid beta-42 independently predicts both late-onset depression and Alzheimer disease. The American Journal of Geriatric Psychiatry. 2010;18(11):973–982.

52. Direk N, Schrijvers EM, de Bruijn RF, et al. Plasma amyloid β, depression, and dementia in community-dwelling elderly. Journal of psychiatric research. 2013;47(4):479–485.

53. Krell Roesch J, Zaniletti I, Syrjanen JA, et al. Plasma derived biomarkers of Alzheimer’s disease and neuropsychiatric symptoms: A community based study. *Alzheimer’s & Dementia: Diagnosis*, Assessment & Disease Monitoring. 2023;15(3):e12461.

54. Liao Y, Chong JR, Kan CN, et al. Plasma NfL, P-tau181, and P-tau181/Aβ42 Ratio in Predicting Mild Behavioral Impairment in Dementia-Free Multiethnic Asian Older Adults With Mixed Pathology in a 5-Year Clinical Cohort. The Journal of clinical psychiatry. 2025;86(1):24m15558.

55. Huang L, Huang Q, Xie F, Guo Q. Neuropsychiatric symptoms in Alzheimer’s continuum and their association with plasma biomarkers. Journal of Affective Disorders. 2024;348:200–206.

56. Steinacker P, Al Shweiki MR, Oeckl P, et al. Glial fibrillary acidic protein as blood biomarker for differential diagnosis and severity of major depressive disorder. Journal of psychiatric research. 2021;144:54–58.

57. Aguzzoli CS, Ferreira PC, Povala G, et al. Neuropsychiatric symptoms and microglial activation in patients with Alzheimer disease. JAMA network open. 2023;6(11):e2345175–e2345175.

58. Miao R, Chen HY, Gill S, et al. Plasma β-amyloid in mild behavioural impairment– neuropsychiatric symptoms on the Alzheimer’s continuum. Journal of geriatric psychiatry and neurology. 2022;35(3):434–441.

59. Naude JP, Gill S, Hu S, et al. Plasma neurofilament light: a marker of neurodegeneration in mild behavioral impairment. Journal of Alzheimer’s disease. 2020;76(3):1017–1027.

60. Leow Y, Wang J, Vipin A, et al. Biomarkers and Cognition Study, Singapore (BIOCIS): Protocol, Study Design, and Preliminary Findings. The Journal of Prevention of Alzheimer’s Disease. Published online 2024:1–13.

61. Petersen RC. Mild cognitive impairment as a diagnostic entity. Journal of internal medicine. 2004;256(3):183–194.

62. Jack Jr CR, Bennett DA, Blennow K, et al. NIA-AA research framework: toward a biological definition of Alzheimer’s disease. Alzheimer’s & dementia. 2018;14(4):535–562.

63. Pendlebury ST, Poole D, Burgess A, Duerden J, Rothwell PM, Oxford Vascular Study. APOE-ε4 genotype and dementia before and after transient ischemic attack and stroke: population-based cohort study. Stroke. 2020;51(3):751–758.

64. Zhong L, Xie YZ, Cao TT, et al. A rapid and cost-effective method for genotyping apolipoprotein E gene polymorphism. Molecular neurodegeneration. 2016;11:1–8.

65. Wilson DH, Rissin DM, Kan CW, et al. The Simoa HD-1 analyzer: a novel fully automated digital immunoassay analyzer with single-molecule sensitivity and multiplexing. Journal of laboratory automation. 2016;21(4):533–547.

66. Llovet JM, Peña CE, Lathia CD, Shan M, Meinhardt G, Bruix J. Plasma biomarkers as predictors of outcome in patients with advanced hepatocellular carcinoma. Clinical Cancer Research. 2012;18(8):2290–2300.

67. Nixon AB, Pang H, Starr MD, et al. Prognostic and predictive blood-based biomarkers in patients with advanced pancreatic cancer: results from CALGB80303 (Alliance). Clinical Cancer Research. 2013;19(24):6957–6966.

68. Babulal GM, Ghoshal N, Head D, et al. Mood changes in cognitively normal older adults are linked to Alzheimer disease biomarker levels. The American journal of geriatric psychiatry. 2016;24(11):1095–1104.

69. Wolfova K, Creese B, Aarsland D, et al. Gender/sex differences in the association of mild behavioral impairment with cognitive aging. Journal of Alzheimer’s Disease. 2022;88(1):345–355.

70. Gao W, Yan X, Yuan J. Neural correlations between cognitive deficits and emotion regulation strategies: understanding emotion dysregulation in depression from the perspective of cognitive control and cognitive biases. Psychoradiology. 2022;2(3):86–99.

71. Hack LM, Tozzi L, Zenteno S, et al. A cognitive biotype of depression and symptoms, behavior measures, neural circuits, and differential treatment outcomes: a Prespecified secondary analysis of a randomized clinical trial. JAMA Network Open. 2023;6(6):e2318411–e2318411.

72. Le Heron C, Holroyd CB, Salamone J, Husain M. Brain mechanisms underlying apathy. *Journal of Neurology*, Neurosurgery & Psychiatry. 2019;90(3):302–312.

73. Rabin JS, Schultz AP, Hedden T, et al. Interactive associations of vascular risk and β-amyloid burden with cognitive decline in clinically normal elderly individuals: findings from the Harvard Aging Brain Study. JAMA neurology. 2018;75(9):1124–1131.

74. Sturm VE, Yokoyama JS, Seeley WW, Kramer JH, Miller BL, Rankin KP. Heightened emotional contagion in mild cognitive impairment and Alzheimer’s disease is associated with temporal lobe degeneration. Proceedings of the National Academy of Sciences. 2013;110(24):9944–9949.

75. Devanand DP, Lee S, Huey ED, Goldberg TE. Associations between neuropsychiatric symptoms and neuropathological diagnoses of Alzheimer disease and related dementias. JAMA psychiatry. 2022;79(4):359–367.

76. Dietlin S, Soto M, Kiyasova V, et al. Neuropsychiatric symptoms and risk of progression to Alzheimer’s disease among mild cognitive impairment subjects. Journal of Alzheimer’s Disease. 2019;70(1):25–34.

77. Guan DX, Rehman T, Nathan S, et al. Neuropsychiatric symptoms: Risk factor or disease marker? A study of structural imaging biomarkers of Alzheimer’s disease and incident cognitive decline. Human Brain Mapping. 2024;45(13):e70016.

78. Lussier FZ, Pascoal TA, Chamoun M, et al. Mild behavioral impairment is associated with β amyloid but not tau or neurodegeneration in cognitively intact elderly individuals. Alzheimer’s & dementia. 2020;16(1):192–199.

79. Piccinni A, Origlia N, Veltri A, et al. Neurodegeneration, β amyloid and mood disorders: state of the art and future perspectives. International journal of geriatric psychiatry. 2013;28(7):661–671.

80. Brown RB. Stress, inflammation, depression, and dementia associated with phosphate toxicity. Molecular Biology Reports. 2020;47(12):9921–9929.

81. Plascencia-Villa G, Perry G. Neuropathologic changes provide insights into key mechanisms of Alzheimer disease and related dementia. The American Journal of Pathology. 2022;192(10):1340–1346.

82. Naude J, Wang M, Leon R, Smith E, Ismail Z. Tau-PET in early cortical Alzheimer brain regions in relation to mild behavioral impairment in older adults with either normal cognition or mild cognitive impairment. Neurobiology of Aging. 2024;138:19–27.

83. Tissot C, Therriault J, Pascoal TA, et al. Association between regional tau pathology and neuropsychiatric symptoms in aging and dementia due to Alzheimer’s disease. Alzheimer’s & Dementia: Translational Research & Clinical Interventions. 2021;7(1):e12154.

84. Walsh DM, Selkoe DJ. Amyloid β-protein and beyond: the path forward in Alzheimer’s disease. Current opinion in neurobiology. 2020;61:116–124.

85. Kim HJ, Park KW, Kim TE, et al. Elevation of the plasma Aβ 40/Aβ 42 ratio as a diagnostic marker of sporadic early-onset Alzheimer’s disease. Journal of Alzheimer’s Disease. 2015;48(4):1043–1050.

86. Janelidze S, Mattsson N, Palmqvist S, et al. Plasma P-tau181 in Alzheimer’s disease: relationship to other biomarkers, differential diagnosis, neuropathology and longitudinal progression to Alzheimer’s dementia. Nature medicine. 2020;26(3):379–386.

87. Karikari TK, Pascoal TA, Ashton NJ, et al. Blood phosphorylated tau 181 as a biomarker for Alzheimer’s disease: a diagnostic performance and prediction modelling study using data from four prospective cohorts. The Lancet Neurology. 2020;19(5):422–433.

88. Mattsson N, Andreasson U, Zetterberg H, Blennow K, Alzheimer’s Disease Neuroimaging Initiative. Association of plasma neurofilament light with neurodegeneration in patients with Alzheimer disease. JAMA neurology. 2017;74(5):557–566.

89. Brandebura AN, Paumier A, Onur TS, Allen NJ. Astrocyte contribution to dysfunction, risk and progression in neurodegenerative disorders. Nature Reviews Neuroscience. 2023;24(1):23–39.

90. Hawksworth J, Fernandez E, Gevaert K. A new generation of AD biomarkers: 2019 to 2021. Ageing research reviews. 2022;79:101654.

91. Simrén J, Elmgren A, Blennow K, Zetterberg H. Fluid biomarkers in Alzheimer’s disease. Advances in clinical chemistry. 2023;112:249–281.

92. Preische O, Schultz SA, Apel A, et al. Serum neurofilament dynamics predicts neurodegeneration and clinical progression in presymptomatic Alzheimer’s disease. Nature medicine. 2019;25(2):277–283.

93. Cicognola C, Janelidze S, Hertze J, et al. Plasma glial fibrillary acidic protein detects Alzheimer pathology and predicts future conversion to Alzheimer dementia in patients with mild cognitive impairment. Alzheimer’s research & therapy. 2021;13:1–9.

94. Spotorno N, Najac C, Stomrud E, et al. Astrocytic function is associated with both amyloid-β and tau pathology in non-demented APOE 4 carriers. Brain Communications. 2022;4(3):fcac135.

95. Pereira JB, Janelidze S, Smith R, et al. Plasma GFAP is an early marker of amyloid-β but not tau pathology in Alzheimer’s disease. Brain. 2021;144(11):3505–3516.

96. Bhalla G, Tanoto P, Vipin A, et al. Current status and future directions for the diagnosis and management of mild cognitive impairment in Southeast Asia: A SEACURE consensus paper. The Journal of Prevention of Alzheimer’s Disease. Published online 2025:100110.

97. Lénárt N, Brough D, Dénes Á. Inflammasomes link vascular disease with neuroinflammation and brain disorders. Journal of Cerebral Blood Flow & Metabolism. 2016;36(10):1668–1685.

98. Janelidze S, Mattsson N, Stomrud E, et al. CSF biomarkers of neuroinflammation and cerebrovascular dysfunction in early Alzheimer disease. Neurology. 2018;91(9):e867–e877.

99. Edwards M, Balldin VH, Hall J, O’Bryant S. Combining select neuropsychological assessment with blood-based biomarkers to detect mild Alzheimer’s disease: a molecular neuropsychology approach. Journal of Alzheimer’s Disease. 2014;42(2):635–640.

100. Petersen M, Hall J, Parsons T, Johnson L, O’Bryant S. Combining select blood-based biomarkers with neuropsychological assessment to detect mild cognitive impairment among Mexican Americans. Journal of Alzheimer’s Disease. 2020;75(3):739–750.

101. Teunissen CE, Verberk IM, Thijssen EH, et al. Blood-based biomarkers for Alzheimer’s disease: towards clinical implementation. The Lancet Neurology. 2022;21(1):66–77.

102. Johansson M, Stomrud E, Lindberg O, et al. Apathy and anxiety are early markers of Alzheimer’s disease. Neurobiology of aging. 2020;85:74–82.

103. Rosenberg PB, Nowrangi MA, Lyketsos CG. Neuropsychiatric symptoms in Alzheimer’s disease: What might be associated brain circuits? Molecular aspects of medicine. 2015;43:25–37.

104. Preman P, Alfonso-Triguero M, Alberdi E, Verkhratsky A, Arranz AM. Astrocytes in Alzheimer’s disease: pathological significance and molecular pathways. Cells. 2021;10(3):540.

105. Rauf A, Badoni H, Abu-Izneid T, et al. Neuroinflammatory markers: key indicators in the pathology of neurodegenerative diseases. Molecules. 2022;27(10):3194.

106. Shen XN, Huang SY, Cui M, et al. Plasma glial fibrillary acidic protein in the Alzheimer disease continuum: relationship to other biomarkers, differential diagnosis, and prediction of clinical progression. Clinical Chemistry. 2023;69(4):411–421.

107. Abdelhak A, Foschi M, Abu-Rumeileh S, et al. Blood GFAP as an emerging biomarker in brain and spinal cord disorders. Nature Reviews Neurology. 2022;18(3):158–172. doi:10.1038/s41582-021-00616-3

108. Heneka MT, Carson MJ, El Khoury J, et al. Neuroinflammation in Alzheimer’s disease. The Lancet Neurology. 2015;14(4):388–405.

109. Orgeta V, Leung P, del-Pino-Casado R, et al. Psychological treatments for depression and anxiety in dementia and mild cognitive impairment. Cochrane Database of Systematic Reviews. 2022;(4).

110. Zhang Y, Chen H, Li R, Sterling K, Song W. Amyloid β-based therapy for Alzheimer’s disease: challenges, successes and future. Signal transduction and targeted therapy. 2023;8(1):248.

